# KAP survey of practicing doctors on antimicrobial stewardship based on openWHO course

**DOI:** 10.1101/2021.03.10.21253238

**Authors:** Simerdeep Kaur, Prativa Sethi, Prasan Kumar Panda

## Abstract

**Background:** The overwhelming, irrational behaviour of using antimicrobial (AM) has added to the amplification and spread of antimicrobial resistance (AMR) burden. Healthcare professionals can curtail the AMR by practicing antimicrobial stewardship (AMS). Keeping this in view WHO has laid down a global action plan to combat AMR including free online availability of openWHO course. So, our study aimed at accessing the knowledge, attitude, and practice (KAP) of practicing doctors towards AMS based on this course in a tertiary care hospital.

**Methods:** The study was conducted among practitioners (faculty, senior residents, junior residents) in different clinical departments. The study was designed as a KAP survey. A validated self-administered questionnaire consisting of 29 questions was designed and shared among 200 participants through the mail and physically. Apart from observing knowledge/attitude/practice gaps, the difference in response to questions was evaluated among various groups (surgeon vs physician, faculty vs senior resident vs junior resident, openWHO course participant vs openWHO course aware non-participant vs openWHO course unaware non-participants.

**Results:** Response rate was 62.5% (n=200). Knowledge on AMS was observed among doctors’ with >50% near correct responses in each question except for the question asking on IV route of AM administration. A significant knowledge gap was found when a comparison was made between faculty members, senior residents, and junior residents (p <0.001) in the spectrum of activity of AM. Almost all the participants agreed that ASP is a necessity in the hospital and believed that ASP reduces healthcare costs and adverse effects of inappropriate AM prescription. A significant difference between the various groups’ aspects was also observed.

**Conclusion:** Knowledge gap on ASP is observed among all HCPs but significant differences among faculty, senior residents, and junior residents, among openWHO course vs unaware openWHO course participant. This shows faculty has to take the lead including more and more practice and education in ASP. The openWHO course may help in achieving this.

## Background

The prevalence of infectious disease ranges from 28.05 to 29.57 per 1000 population, which is very high in a developing country like India.^1^ Antimicrobial (AM) agents have played a critical role in reducing the burden of communicable diseases across the world with a good reason that many have considered them ‘wonder drugs’.^2^ However, AM consumption too increases. According to a study published in the proceedings of the National Academy of Sciences, AM consumption in India has increased to 65%.^3^ This increased use leads to the emergence of antimicrobial resistance (AMR) that is creating ‘superbugs’ that make treating basic infections difficult and AMR is one of the biggest threats to global health, food security, and development today.^2,4^

Antimicrobial resistance develops over time, usually through genetic changes when microorganisms are exposed to AM drugs.^4^ Two main contributing factors - excessive use of AM and inadequate dose of AM have led to the ramification of resistant organisms. New resistance mechanisms are emerging and spreading globally, threatening our ability to treat common infectious diseases. The cost of healthcare for patients with resistant infections is becoming higher than care for patients with non-resistant infections due to longer duration of illness, ICU stays, additional tests, and the use of more expensive drugs.^5-7^ The third GLASS report presents the frequency of AMR in 2,164,568 patients with laboratory-confirmed infections in 66 countries, territories, and areas in 2018. The rate of resistance to ciprofloxacin commonly used to treat urinary tract infections, varied from 8.4% to 92.9% for *E. coli* and from 4.1% to 79.4% for *K. pneumonia* respectively.^8^ In 2013, CDC published the first AR threat report, which rang the alarm for an overwhelming increase in AMR.^9,10^ There has been a 28% decrease in death from AMR in hospitals since the 2013 report.^11^Furthermore, lack of new antibiotics threatens global efforts to contain drug-resistant infections.^10,12^

What is the solution? Antimicrobial stewardship (AMS) is a coherent set of actions that promote the responsible use of AM.^13^ An antimicrobial stewardship program (ASP) is an organizational or system-wide health-care strategy to promote the appropriate use of AM through the implementation of evidence-based interventions.^14^ A broad range of interventions has been implemented to improve ASP, e.g. TARGET toolkit in the UK, openWHO course.^15,16^ Till now many knowledge, attitude, practice (KAP) studies on AMR have been conducted among community members, medical undergraduate students, which has shown AMR is an increasing national problem, and the attitude of self-medication in about 46% of participants, and the need for more educational tools in non-medical professionals.^17-23^ All studies have shown AMR being a great problem (>90%) to public health as well as a national problem, but <60% rate to be a problem in their real clinical practice. Very few, less than 30% knew the prevalence of multi-drug resistant in their hospitals. Few KAP studies have been done among doctors in tertiary care centres in India.^24,25^ In all these studies the authors have identified gaps of doctors towards AMR, which are important to promote the rational use of AM and to develop their hospital ASP, however, no one utilizes any specific guideline or online course content if they are practicing w.r.t. AMS/ASP.

This study aims to determine the knowledge, attitude, and practice among doctors towards AMS as per onlineWHO course and to look for the impact of free openWHO course on AMS. we hope that a study like this also improves the KAP of antimicrobials among doctors, the ultimate goal being the control of AMR.

To report our findings, we followed the STROBE (Strengthening the Reporting of Observational Studies in Epidemiology) guideline.

## Materials and methods

### Study setting and design

We conducted a hospital-based observational cross-sectional study among doctors of different departments in a tertiary care hospital, AIIMS RISHIKESH from July to September 2019 over 2 months. All India institute of medical science (AIIMS) is one of the centre of excellence harbouring 1100 beds acting as referral centres from many places in north India.

### Study population

Doctors from different departments (medicine, surgery, paediatrics, obstetrics & gynaecology, etc) of AIIMS Rishikesh having expertise in their field, were recruited. As these departments were having maximum direct exposure and use of AM to the patients, a total of 200 doctors were recruited in the study. The sample size was calculated by taking KAP prevalence in a similar study done before. Doctors included faculties, senior residents, and junior residents from 2^nd^ and 3rd-year post-graduate courses, who were mainly deciding the patient treatment.

### Assessment material

The questionnaire is self-structured after searching medical literature for comparable studies and adapting questions based on the online openWHO course: Antimicrobial stewardship: A competency-based approach (freely available).(15) The questionnaire was verified and authenticated by subject experts for its contents and relevance. Pre-validation of the questionnaire was also done, resulting in a total of 27 questions (having subparts).

The questionnaire included four sections. Section one started with the characterization of practicing doctors’ professional profiles (staff position, the field of specialty, and years of experience) in this tertiary care hospital setting and whether or not they have participated in a free online openWHO course (by collecting the participant certificate). The second section of the questionnaire assessed doctors’ knowledge on AMS (pharmacokinetics of AM, basics and components of AMS, mechanism on AMR, route of AM) using 10 questions (some having 4-5 subparts). The third section was divided into two subparts to assess doctors’ attitudes towards AMS and the openWHO course. The last section had a total of six questions out of which four questions assessed doctors’ practices and decision-making skills in a particularly common case scenario, one question was used to assess their daily clinical practice of AM use, and the last question assessed surgeons’ practice skills to prevent skin and soft tissue infection based on WHO guideline.

The questions were evaluated on a 5-point Likert scale with response options of Strongly Disagree/Disagree/Neither agree nor disagree/Agree/Strongly agree.

### Operational definitions

The following definitions were used to select the participants enrolled in the study:

- Target population: All the practicing doctors in a tertiary health care setting
- Residents: Junior Residents and Senior Residents
- Faculty: Consultant (Assistant, Associate, Additional Professors, and Professors)
- Participants: The members of the target population present at the time of conducting the study who gave consent were defined as Participants
- Responders: The participants who return the filled questionnaire to enrol in the study were defined as the Responders
- Non-responder: Failing to fill the questionnaire after 3 subsequent visit
- Lost to follow-up: Those responders who did not return the questionnaire after 3 repeated reminders if they filled form incompletely at initial interaction

### Methodology

The study was assessed and approved by the institutional ethical committee before starting the survey. Members of the target population who were present within the period of the survey and gave consent by filling the google form shared to them by email and or WhatsApp were included in the study. The questionnaire was distributed to working doctors of all clinical departments who prescribes antimicrobials. Doctors from departments of Radiology, Anaesthesia, Psychiatry, and Physical Medical Rehabilitation were excluded due to the rarity of antimicrobials use. Also, the Interns working in the hospital were excluded. Participants were visited during working hours and were given hard copies or online survey links of the questionnaire according to the participant’s choice. Those who had not submitted in the first visit were re-visited or re-mailed up to three times till compliance was established after which it was decided non-responder. No incentives were offered for participation. Availability of drug and therapeutic committee and hospital antibiotic policy was there.

### Data Analysis

After collecting the questionnaire and obtaining the required data in Microsoft Excel® sheet, they were evaluated for completeness. All gathered data grouped into surgeon vs physician, faculty vs senior resident vs junior resident, and open WHO participant vs aware non-participant vs unaware non-participant. Participants were also analysed in a subgroup based on the duration of medical experience after MBBS. Data analysis was done using the Statistical Package for Social Sciences (SPSS® 24.0, USA) and interpreted. Proportions were calculated. Pearson Chi-square test was used for categorical data. The final results were compared with the right answers for the questions, and tables and diagrams were used to present the results. Differences amongst groups were tested using the Chi-squared test. P-value <0.05 was considered statistically significant.

## Results

### Basic characteristics

The questionnaire was shared among 200 participants through their ease of convenience, either online (mail) or offline, 125 participants (response rate 62.5%) from sixteen departments completed the study and were analysed under categories of departments, positions, years of practice, and user profile of openWHO course (Fig. 1). The data showed maximum participation from the Department of Medicine and most of the participants (77.6%) were non-participant of the openWHO course. On asking the reason for the same, most of them answer about the lack of information.

**Figure 1:**
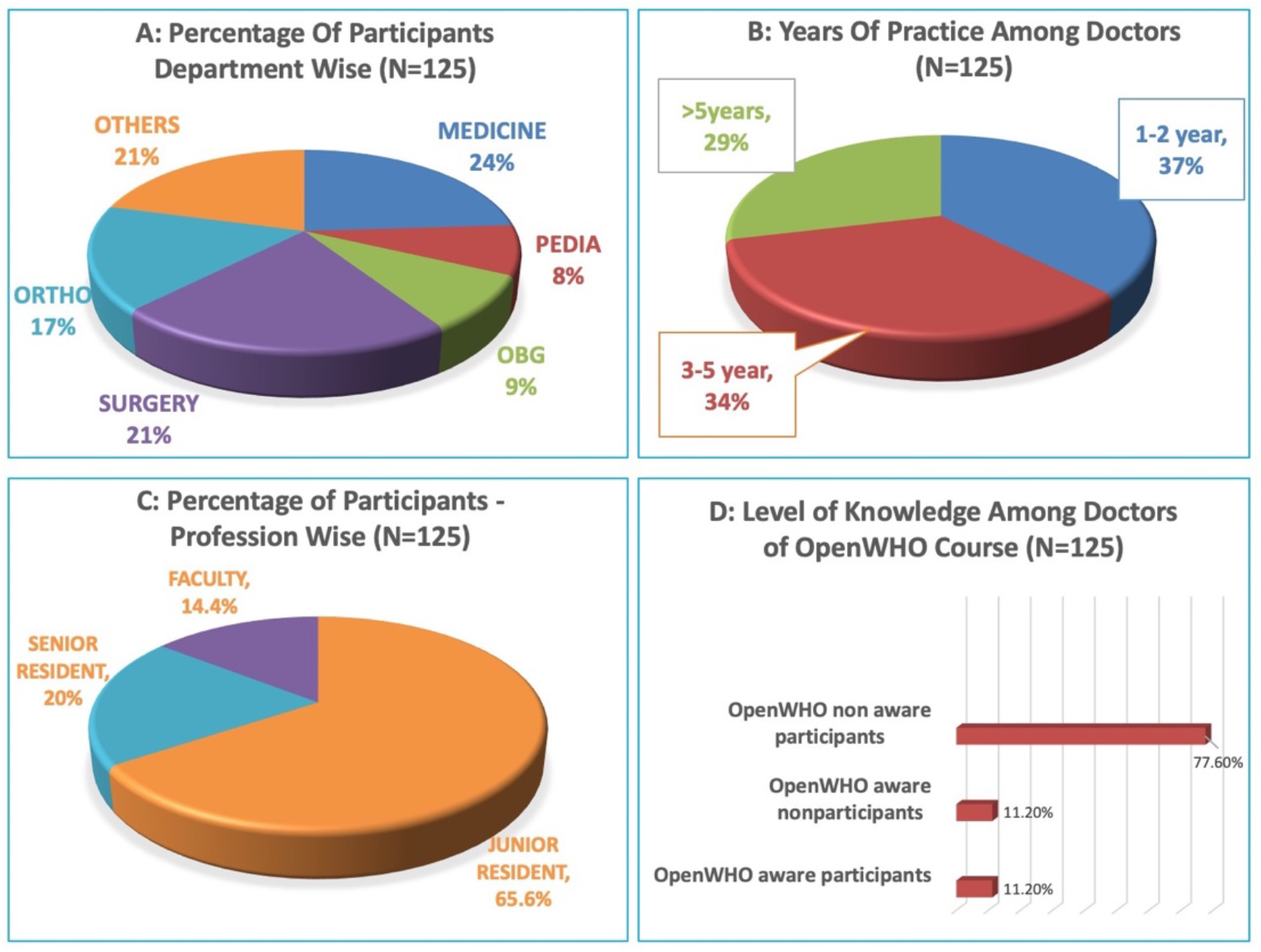
Basic characteristics of participants w.r.t. various sub-groups.

### Knowledge about antimicrobial stewardship

After analysing the responses from the knowledge section, it was found that almost all participants considered a correct diagnosis, dose, and duration as an important principle of AMS. However, 24% (Strongly disagree=9.6%, Disagree=14.4%) of doctors did not consider the route of administration of the drug as an AMS principle and 30.4% (n=38) had no opinion for the given statement (Fig. 2). Less than fifty percent of doctors’ (48.89%) didn’t favour using the broadest spectrum of antimicrobials (D= 33.6%, SD = 15.29%) at the initial and 67.2 % were against using broad-spectrum antimicrobials irrespective of the severity of infection (SD=28%, D=39.2%). The great majority 59.9% (SA= 15.5%, A = 44.4%) doctors’ agreed to the fact that the “Emergence of AMR is inevitable.” While assessing doctors’ knowledge on AMR mechanism, 34.4% of doctors consider increased influx of drug into the bacterial cell as one of the mechanisms by which micro-organisms acquire resistance. In an antimicrobial with concentration-dependent killing, more than fifty percent of doctors’ (SA=16%, A=43.2%) consider large infrequent dosing as well as optimization of AM duration with a concentration over MIC (SA=27.2%, A=28%) as an appropriate regimen. On questioning about the intervention types of AMS, the majority (64%, 65%) don’t know about pre-authorization and formulary restriction respectively.

**Figure 2:**
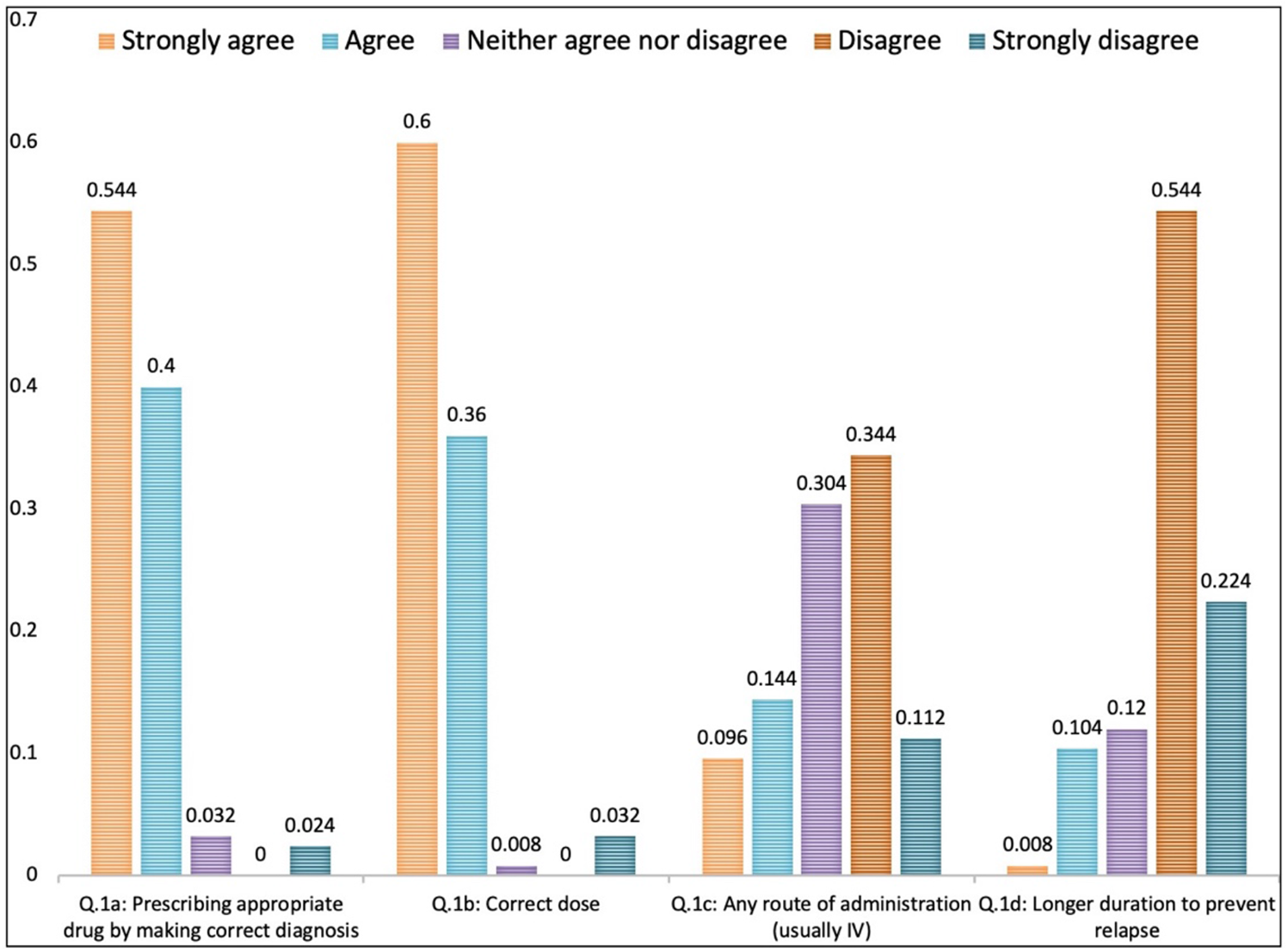
Responses to questions on knowledge of basic principles of AMS.

### Comparison of knowledge

As shown in table 1, there was a significant knowledge gap in considering ‘appropriate dosage to site and type of infection’ as an AMS component (p=0.002) and inactivation of AM as a major mechanism of resistance (p=0.004). There was not much significance in knowledge between surgeons and physicians except in four questions. Only six surgeons in comparison to eleven physicians’ strongly disagreed with the use of broad-spectrum AM (p=0.020).

**Table 1:**
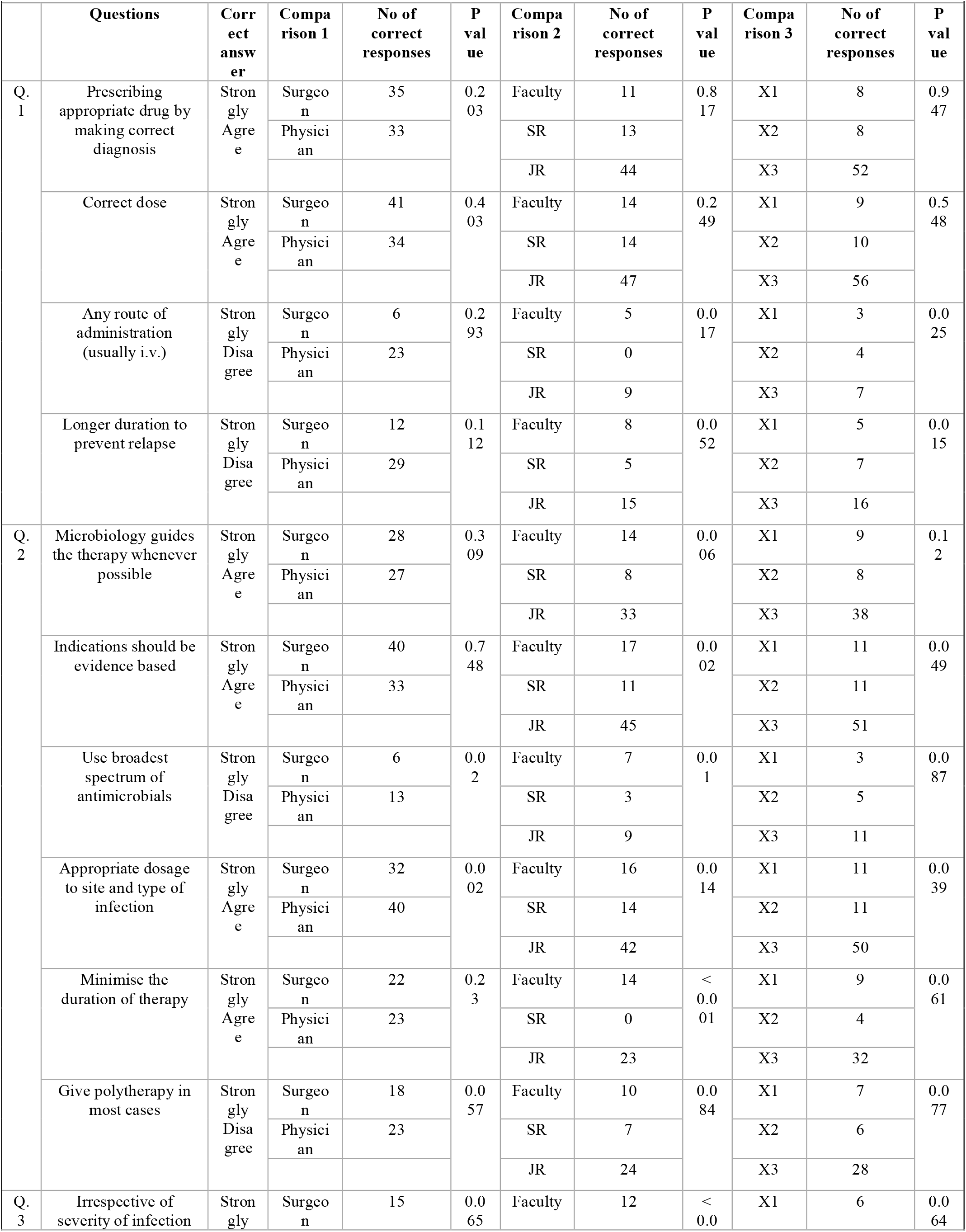
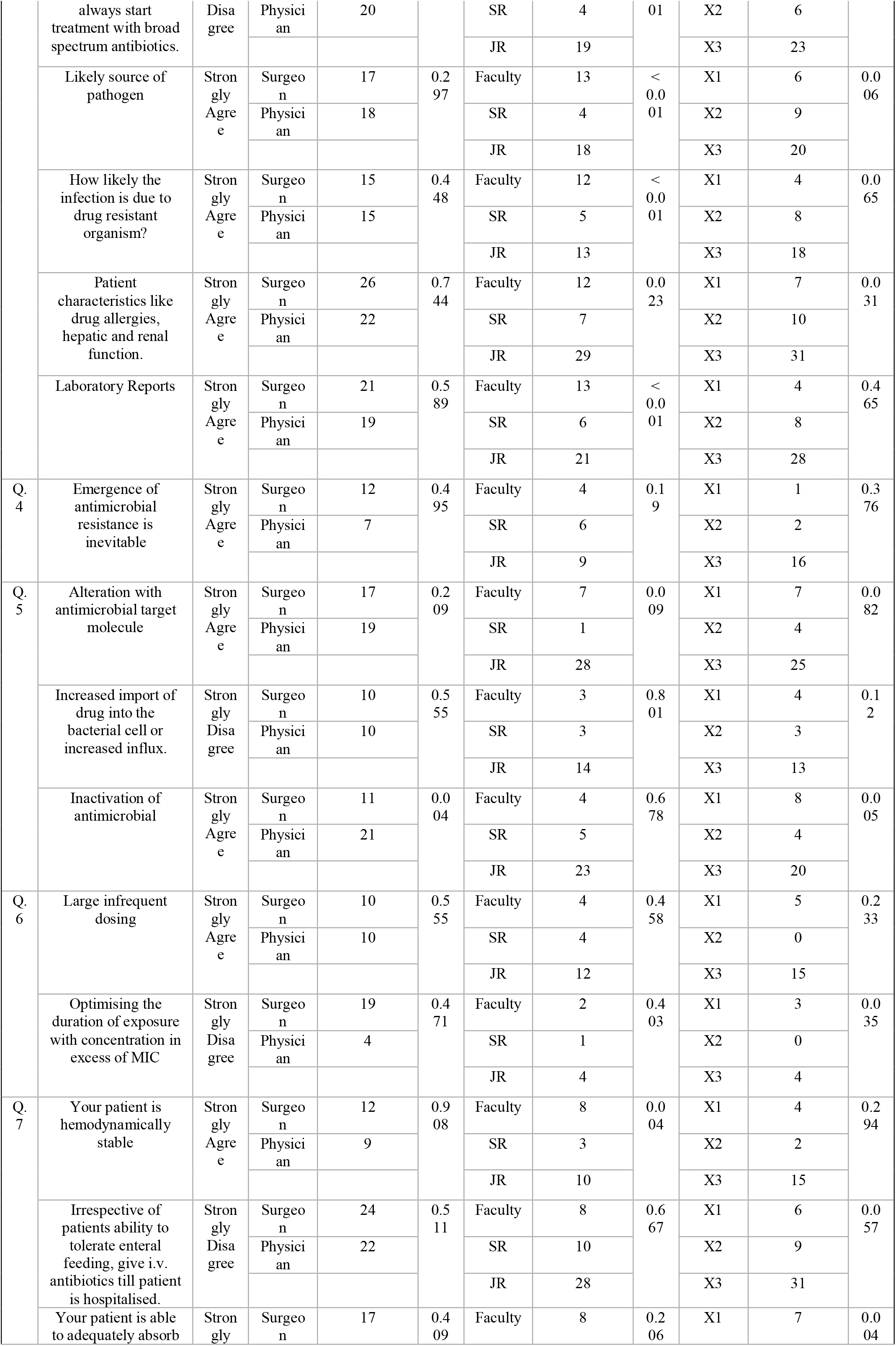
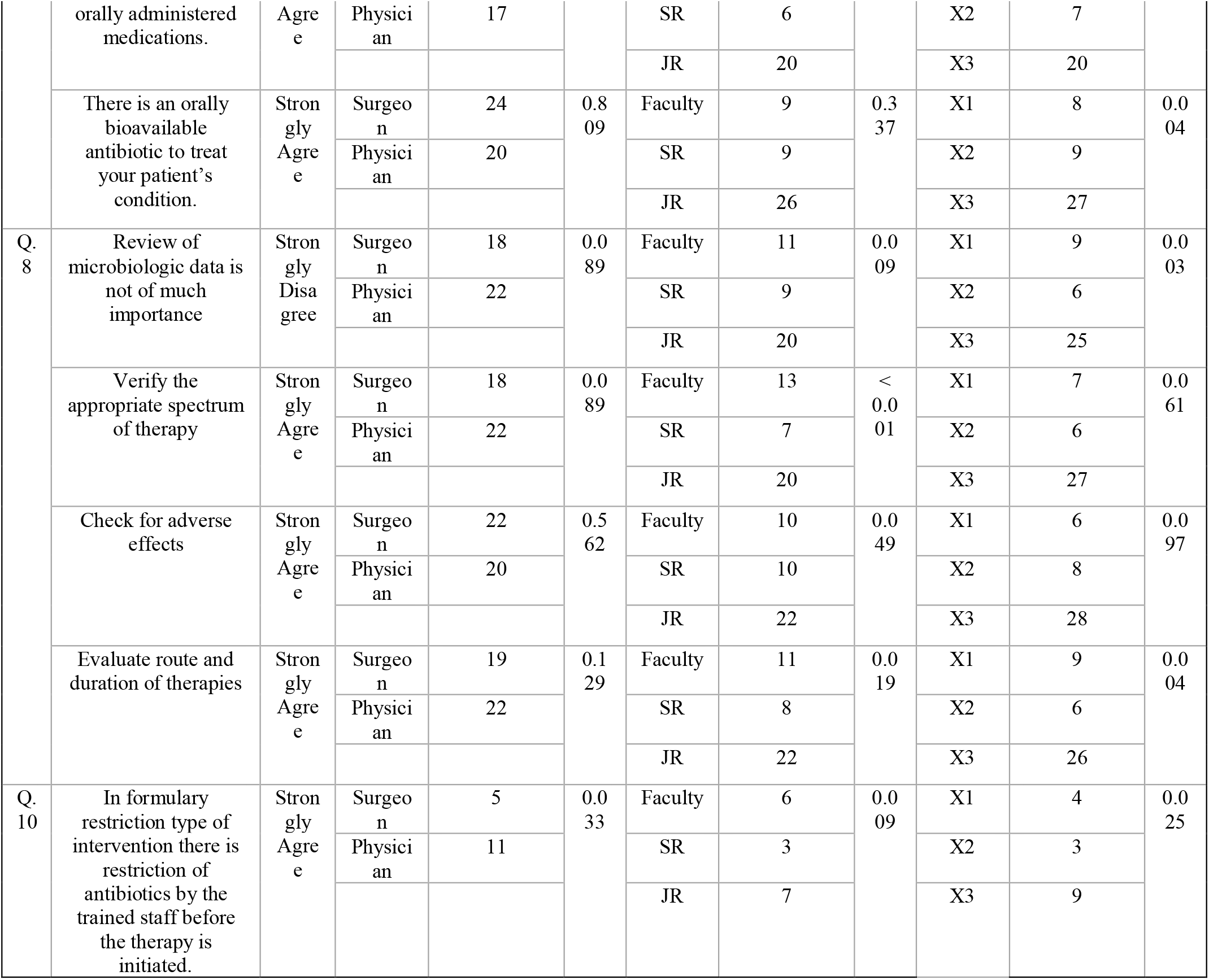
Comparison of Knowledge Among Surgeon Vs Physician, Faculty Vs Senior Resident (SR) Vs Junior Resident (JR), OpenWHO Participants (X1) Vs Aware Non-participants (X2) Vs Unaware Non-participants (X3)

Comparison of knowledge among faculty, senior resident (SR), and junior resident (JR), showed a significant knowledge gap between them with faculty members giving a maximum number of correct answers followed by SRs and JRs which gave a different number of correct answers in different questions. Maximum significance was seen in questions asking factors to decide the spectrum of AM therapy (p<0.001). This indirectly shows the year of experience in clinical practice during which they would have treated the resistant organism.

### Attitude habits towards AMS

Almost all the participants agreed that ASP is a necessity in their hospital as well as it reduces healthcare cost and adverse effects of inappropriate AM prescription (Fig. 3).

**Figure 3:**
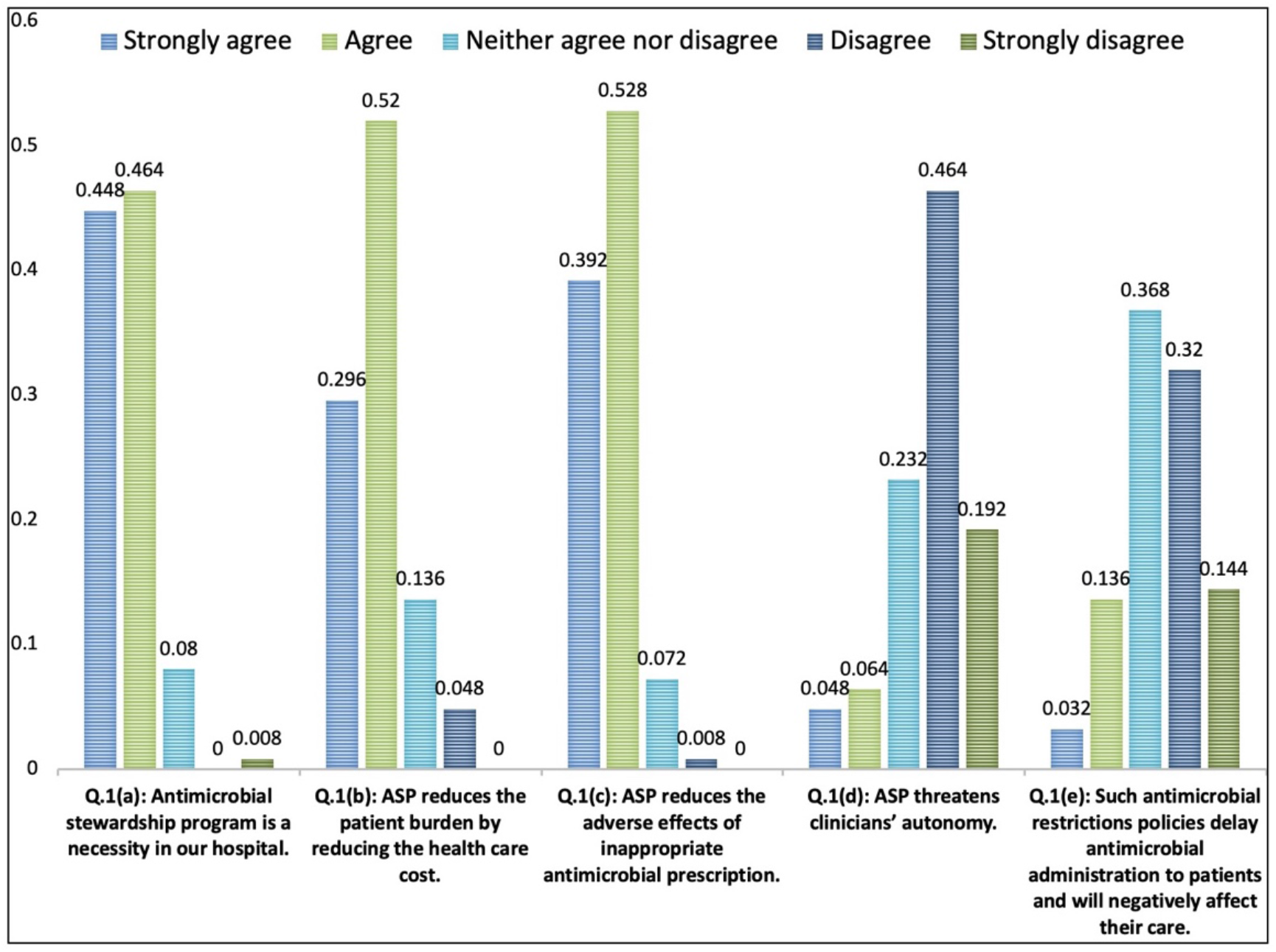
Responses to questionnaire on attitude towards antimicrobial stewardship program.

The majority of participants who had participated in the openWHO course towards AMS agreed on the course to be made compulsory for all HCW (Fig. 4). Similarly, the majority preferred to take the course than seeking advice from the seniors. But almost half of the participants thought that the treatment options are not too ideal to be implicated in the daily practice.

**Figure 4:**
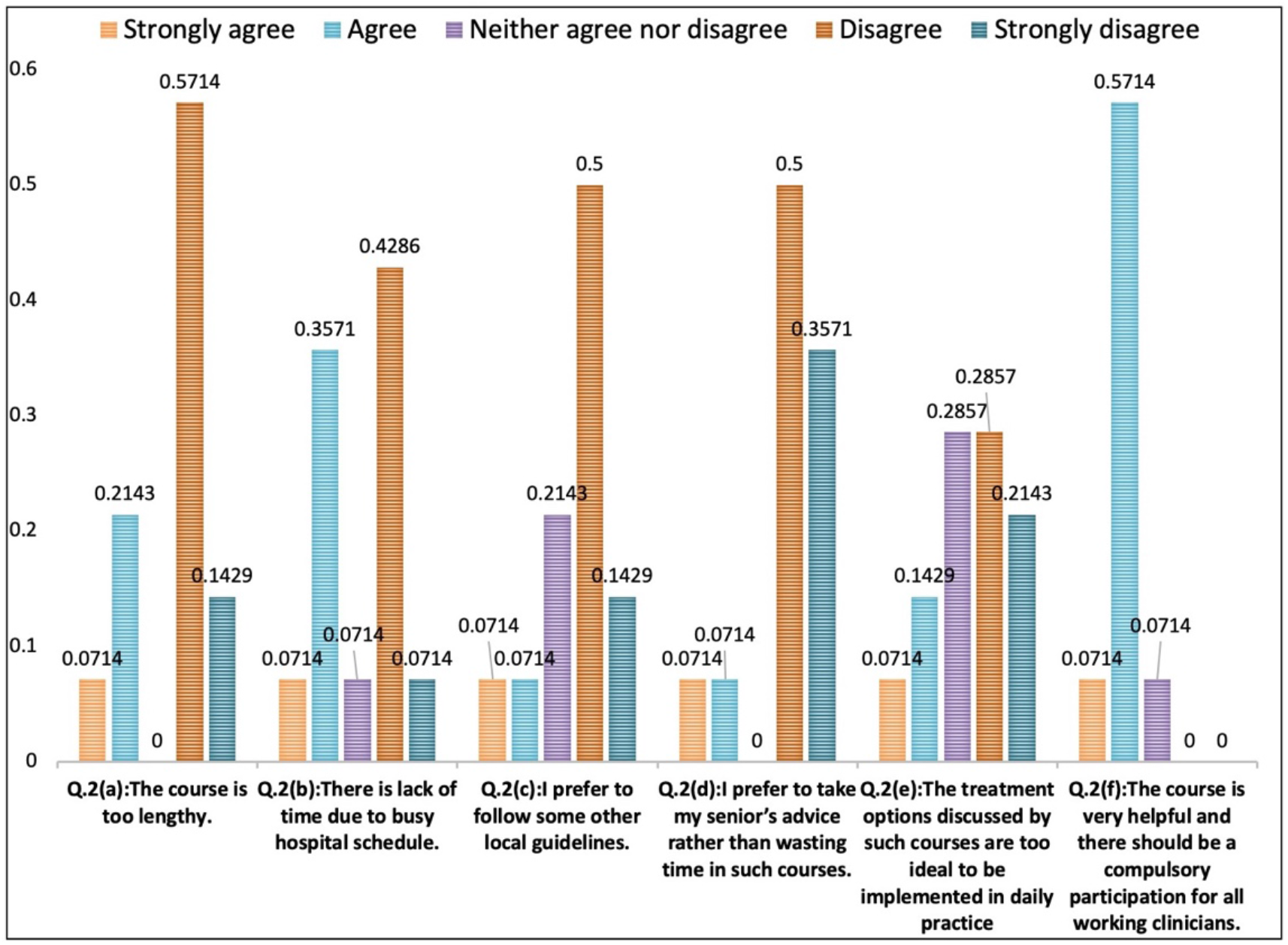
Responses to questionnaire on attitude towards AMS by openWHO participants.

### Comparison of attitude

The study didn’t show any significant differences between surgeons and physicians in attitude habits (Table 2). But there were significant differences among faculty and senior resident in the attitude habit, on health care cost reduction by implementing AMS (p<0.004). There were significant differences in attitude habit among openWHO aware participants than openWHO unaware participants (p<0.05)

**Table 2:**
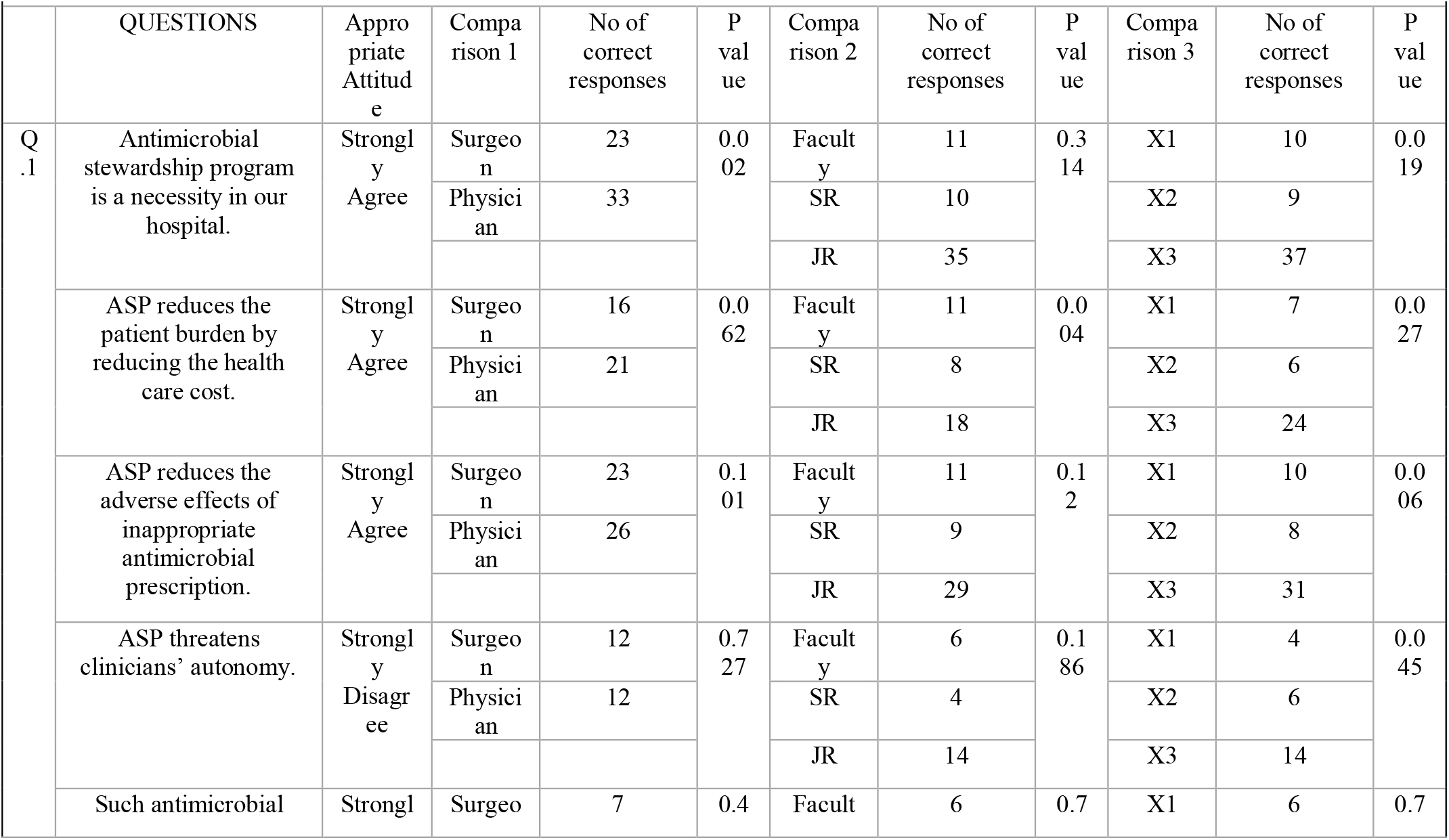
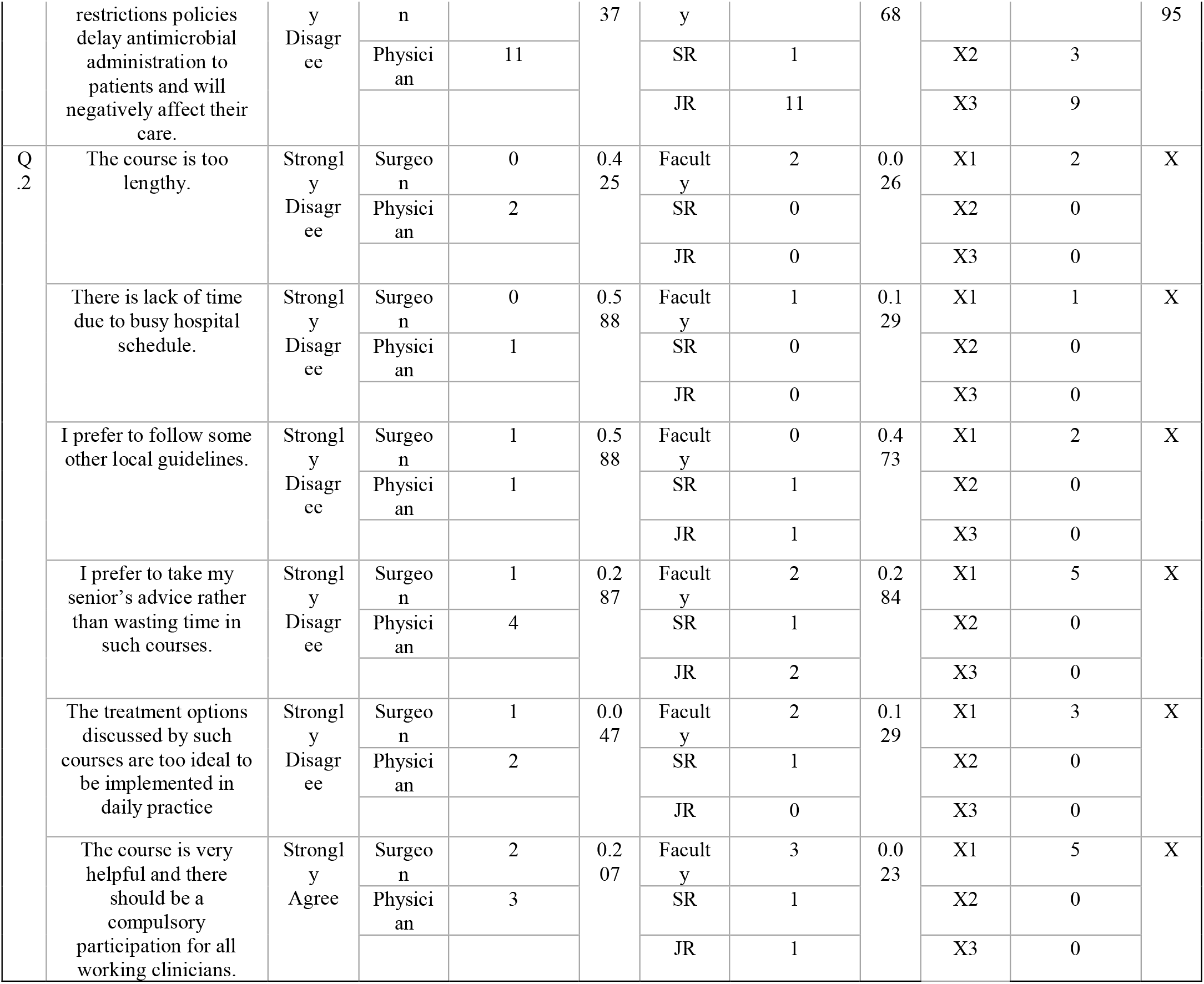
Comparison of Attitude Among Surgeon Vs Physician, Faculty Vs Senior Resident (SR) Vs Junior Resident (JR), OpenWHO Participants (X1) Vs Aware Non-participants (X2) Vs Unaware Non-participants (X3)

|  |  |  |  |  |  |  |  |  |  |  |  |
| --- | --- | --- | --- | --- | --- | --- | --- | --- | --- | --- | --- |
|  | QUESTIONS | Appropriate Attitude | Comparison 1 | No of correct responses | P value | Comparison 2 | No of correct responses | P value | Comparison 3 | No of correct responses | P value |
| Q. 1 | Antimicrobial stewardship program is a necessity in our hospital. | Strongly Agree | Surgeon | 23 | 0.002 | Faculty | 11 | 0.314 | X1 | 10 | 0.019 |
|  |  |  | Physician | 33 |  | SR | 10 |  | X2 | 9 |  |
|  |  |  |  |  |  | JR | 35 |  | X3 | 37 |  |
|  | ASP reduces the patient burden by reducing the health care cost. | Strongly Agree | Surgeon | 16 | 0.062 | Faculty | 11 | 0.004 | X1 | 7 | 0.027 |
|  |  |  | Physician | 21 |  | SR | 8 |  | X2 | 6 |  |
|  |  |  |  |  |  | JR | 18 |  | X3 | 24 |  |
|  | ASP reduces the adverse effects of inappropriate antimicrobial prescription. | Strongly Agree | Surgeon | 23 | 0.101 | Faculty | 11 | 0.12 | X1 | 10 | 0.006 |
|  |  |  | Physician | 26 |  | SR | 9 |  | X2 | 8 |  |
|  |  |  |  |  |  | JR | 29 |  | X3 | 31 |  |
|  | ASP threatens clinicians' autonomy. | Strongly Disagree | Surgeon | 12 | 0.727 | Faculty | 6 | 0.186 | X1 | 4 | 0.045 |
|  |  |  | Physician | 12 |  | SR | 4 |  | X2 | 6 |  |
|  |  |  |  |  |  | JR | 14 |  | X3 | 14 |  |
|  | Such antimicrobial | Strongly | Surgeon | 7 | 0.4 | Faculty | 6 | 0.7 | X1 | 6 | 0.7 |
|  | restrictions policies delay antimicrobial administration to patients and will negatively affect their care. | y<br>Disagree | n<br>Physician | 11 | 37 | y<br>SR | 1 | 68 | X2 | 3 | 95 |
|  |  |  |  |  |  | JR | 11 |  | X3 | 9 |  |
| Q.2 | The course is too lengthy. | Strongly Disagree | Surgeon | 0 | 0.425 | Faculty | 2 | 0.026 | X1 | 2 | X |
|  |  |  | Physician | 2 |  | SR | 0 |  | X2 | 0 |  |
|  |  |  |  |  |  | JR | 0 |  | X3 | 0 |  |
|  | There is lack of time due to busy hospital schedule. | Strongly Disagree | Surgeon | 0 | 0.588 | Faculty | 1 | 0.129 | X1 | 1 | X |
|  |  |  | Physician | 1 |  | SR | 0 |  | X2 | 0 |  |
|  |  |  |  |  |  | JR | 0 |  | X3 | 0 |  |
|  | I prefer to follow some other local guidelines. | Strongly Disagree | Surgeon | 1 | 0.588 | Faculty | 0 | 0.473 | X1 | 2 | X |
|  |  |  | Physician | 1 |  | SR | 1 |  | X2 | 0 |  |
|  |  |  |  |  |  | JR | 1 |  | X3 | 0 |  |
|  | I prefer to take my senior's advice rather than wasting time in such courses. | Strongly Disagree | Surgeon | 1 | 0.287 | Faculty | 2 | 0.284 | X1 | 5 | X |
|  |  |  | Physician | 4 |  | SR | 1 |  | X2 | 0 |  |
|  |  |  |  |  |  | JR | 2 |  | X3 | 0 |  |
|  | The treatment options discussed by such courses are too ideal to be implemented in daily practice | Strongly Disagree | Surgeon | 1 | 0.047 | Faculty | 2 | 0.129 | X1 | 3 | X |
|  |  |  | Physician | 2 |  | SR | 1 |  | X2 | 0 |  |
|  |  |  |  |  |  | JR | 0 |  | X3 | 0 |  |
|  | The course is very helpful and there should be a compulsory participation for all working clinicians. | Strongly Agree | Surgeon | 2 | 0.207 | Faculty | 3 | 0.023 | X1 | 5 | X |
|  |  |  | Physician | 3 |  | SR | 1 |  | X2 | 0 |  |
|  |  |  |  |  |  | JR | 1 |  | X3 | 0 |  |

### Practice assessment of participants

The detailed responses of all the participants were noted (Table 3). Nearly 79.2% of participants reviewed AM at regular intervals. 40.8% of doctors routinely prescribed AM in suspected infections. Only 16.8% of doctors’ were guided by microbiologists while 88.8% of them were guided by pharmacologists in their daily clinical practice. Most of the doctors’ performed poorly when questions were asked based on a clinical scenario.

**Table 3:**
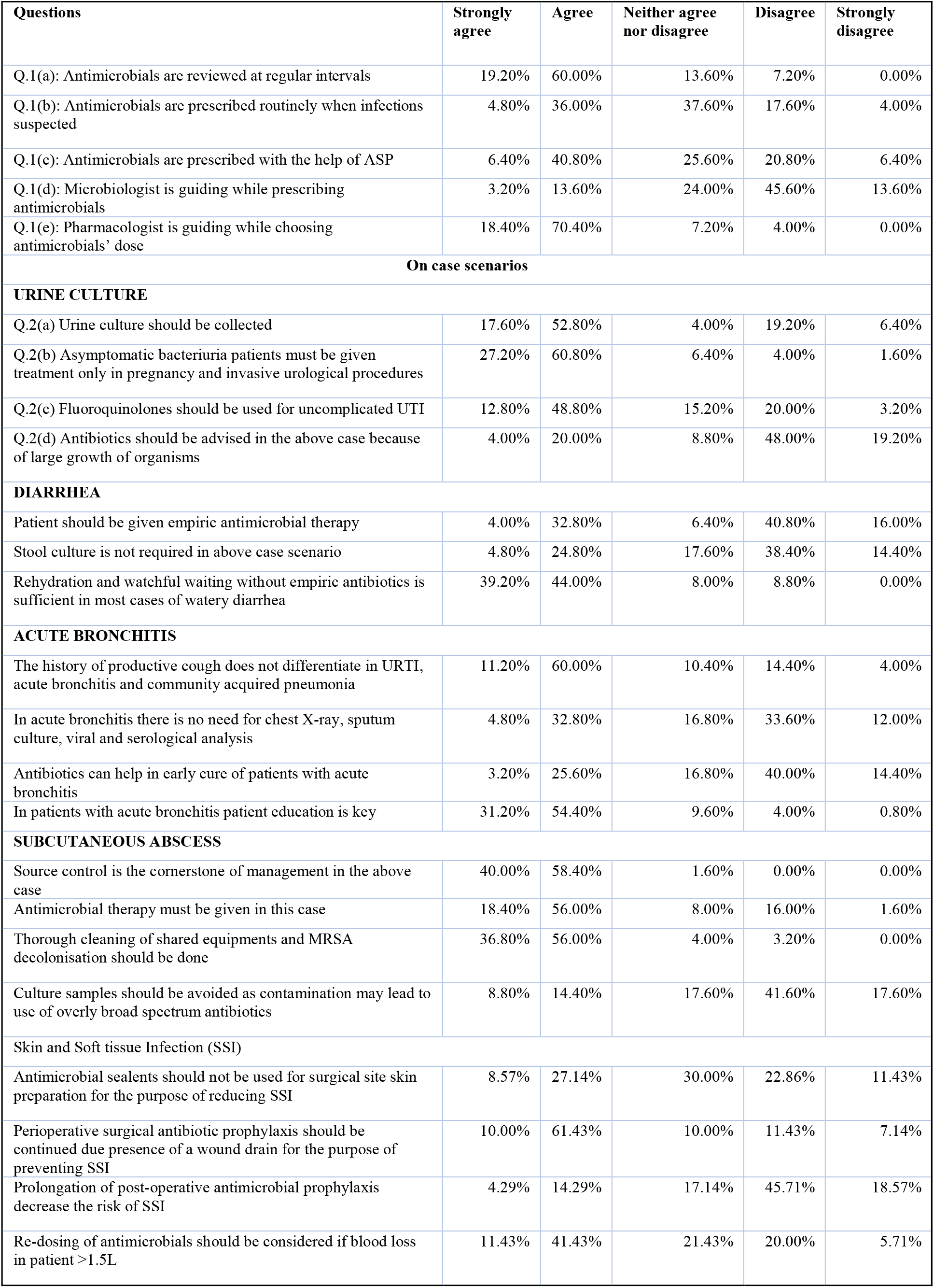
Practice habits of participants towards antimicrobial stewardship.

### Comparison of practice habits

No significant differences were found in practice habits of surgeons and physicians’ except in one question asking about the use of fluoroquinolones in uncomplicated urinary tract infections in which surgeons lag behind physicians’ (p=0.022) (Table 4). Similarly, a significant difference in practice habits among faculty members, SRs, and JRs was found in: need to collect a urine sample in asymptomatic female (p=0.011), the significance of history to differentiate between community-acquired pneumonia, acute bronchitis, and URTI (p=0.006), and prolongation of post-operative AM prophylaxis to reduce surgical site infection (p<0.001). All three questions were performed better by faculty members as seen in the knowledge section. OpenWHO Participants performed better than aware non-participants and unaware non-participants with a significant difference in only two questions asking the use of AM in asymptomatic bacteriuria in pregnant patients (p=0.049) and the use of fluoroquinolones in uncomplicated UTI (p=0.006).

**Table 4:**
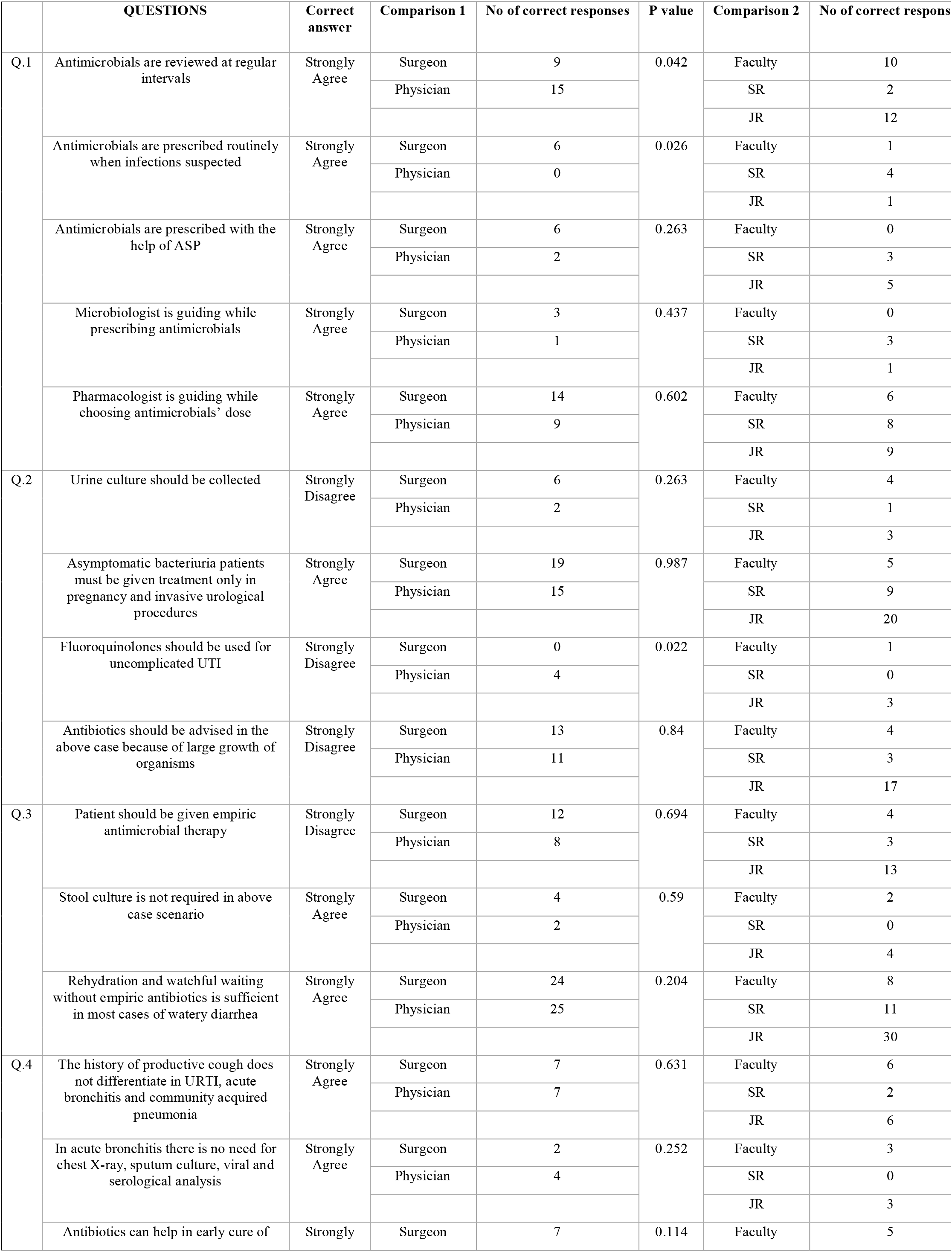
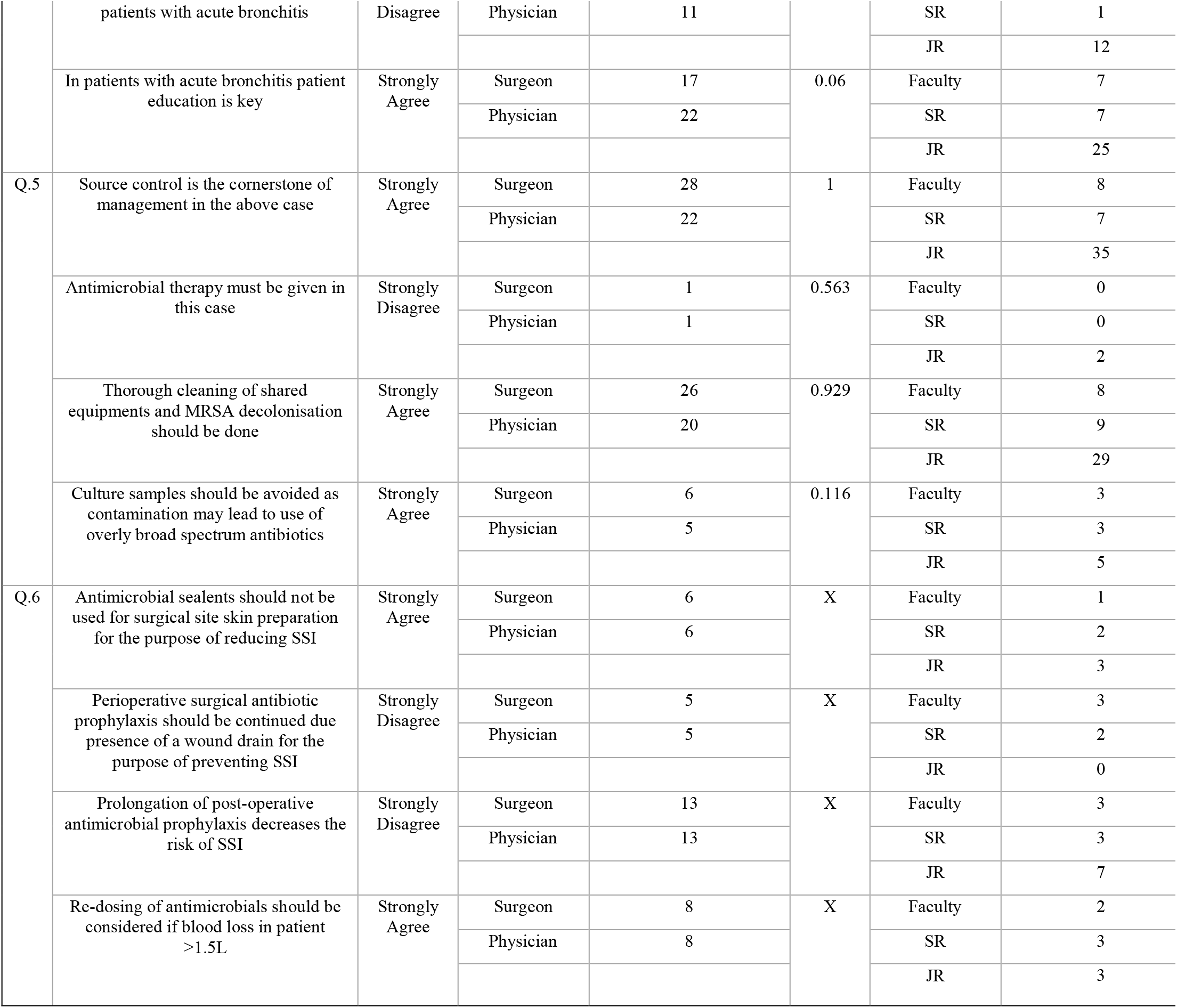
Comparison of Practice Among Surgeon Vs Physician, Faculty Vs Senior Resident (SR) Vs Junior Resident (JR), OpenWHO Participants (X1) V.

## Discussion

Antimicrobial resistance (AMR) created as a ‘superbug’ now has become the most worrisome issue in health care set-up. The perfect knowledge of antimicrobials’ spectrum of activity and its relation with resistance can help health care workers in the correct use of antimicrobials and therefore reducing AMR.^26^ To combat the same WHO has initiated the openWHO online platform: “Antimicrobial stewardship: a competency-based approach” a basic free course having fundamentals on AMS and can be a better way to assess doctors’ from a wide variety of specialties, with variable experience profiles regarding AM in their clinical practice.^13^ Therefore, in this cross-sectional study, we decided to evaluate the KAP of practicing doctors based on this openWHO course on AMS. The study results were quite astonishing, that very few residents and faculty are aware of the availability of such course and quite less participation in this online platform those who were aware. However, the study showed a positive impact of knowledge and experience of faculty members on AMS than the senior and junior residents. It was also proved that openWHO participants gave correct answers in managing the real-world clinical scenario than nonparticipants. To the best of our knowledge, this represents the only study investigating these factors while considering the impact of a massive free online openWHO course in an Indian tertiary care setup.

A study conducted by Byrne et al, on the fact of ‘overuse and misuse of antibiotic: drivers behind consumer behavior amongst the general population’ shows 74% of the individual is on AM for the last year.^27^ In our study too, 19.2 % of the doctors agreed-upon starting broad-spectrum antimicrobials irrespective of the severity of the infection. A large percentage of doctors did not consider the route of administration as an AMS principle (24%) which represents a lack of basic knowledge of AMS in tertiary care doctors’ too. So this high burden of use of AM can only be rationalized by thorough knowledge of AMS and its basic principle - timeliness, appropriateness, adequacy, route, and duration of AM usage.

In our study, fair theoretical knowledge was seen among doctors’. More than fifty percent near correct responses in each basic question on principles of AMS were obtained except for the question asking ‘IV route of AM administration is most commonly preferred irrespective of the severity of infection’ which was strongly disagreed by 11.2%. This has been similar in many studies with fair theoretical knowledge among doctors on AM use.^24,26,28^ This must-have resulted in higher use of IV uses of AM in the hospital. This hints at the early practice of low-hanging fruits of ASP like IV to oral switch, STOP order.

A sufficient knowledge gap was also seen in questions asking about the use appropriate dosing regimen of AM having concentration-dependent killing. Similar results have been seen from the KAP study of Fathi et al that showed the question regarding dose adjustments had the highest frequency of wrong answers of 40% vs 43% in our study.^29^ This indicates there is an urgent unmet need (maybe mandatory) for better training for residents and faculties on this topic in the curriculum. This will result in good clinical practices and avoid inappropriate AM dosing. In few tertiary care institutions in India, it is already a foundation course for residents before appearing in the final master’s degree examination.

As mentioned in figure 1, participation in the openWHO course is very low among doctors. A study conducted by Ghosh A et al showed that 12.5% of doctors had attended a training program in the last 1 year, though it was not based on the openWHO course, still, it gives an impression of the very low percentage of doctors are aware as well as seek to participate in such academic/knowledge update curriculum.^24^ This is highly unacceptable since the course is freely available and in this tertiary care hospital, it was mandatory to be certified with this course by the director 1year before this study initiation. Hence, human behaviour of less upto date needs to be analysed and immediate preventive actions to be ascertained.

In the present study, the participants showed a better practice attitude towards common illness than similar studies done before. A study by Ghosh A et al showed 46.87% of doctors didn’t prescribe AM for simple URTI compared to 71.2% of doctors in our study.^24^ In our study, 56.8% of doctors were against using AM in uncomplicated diarrhoea compared to 59.38% of doctors in the above study, which is comparable. However, to be 100% compliant with AMS, we need better practice attitudes ahead then only AMR can be prevented in toto. There is not much difference between surgical and non-surgical participants, in the knowledge, attitude, and practice survey of this study except in questions concerning appropriate dosage to site and type of infection and promoting the use of broad-spectrum antibiotics. Hence surgeon needs to focus training on the right dose and spectrum of AM to consider. However, there is a significant difference among faculty, senior resident, and junior residents, which supports higher years of experience and knowledge in faculty members. Faculties outperform in various questions in knowledge and practice sections involving deciding AM based on route and duration. Faculty were better able to make decisions when to use broad-spectrum AM. There were significant attitude differences among faculty and senior resident on health care cost reduction by implementing ASP, while a similar study was done comparing junior and senior doctors have shown significant attitude and practice differences in the resistant organism.^18,25^ This emphasizes that the treatment decision must be taken by the faculty. And faculty should be prima facie while AM is chosen, not by free-hand residents in most Indian hospitals.

This study for the first time evaluated the AMS based on openWHO course and compared the KAP survey in aware and unaware non-participants. As the total participant were only 11.2%, the study didn’t show the actual result, so can’t be generalised to the whole population of doctors. In another way, the study reflected true KAP results without any theoretical biases. Being a KAP study, the number of participants, selection bias due to the nature of the KAP study, and involvement of self-volunteering in answering the questions can be mentioned. A limitation of KAP studies is the probability that participants may give socially desirable answers rather than their actual beliefs. Studies taking place in teaching hospitals can be more prone to this limitation.

## Conclusion

To conclude, the present KAP survey has generated information about the AMS approach of medical doctors from a tertiary care teaching hospital in North India. It brings a new comparison group of openWHO aware vs unaware groups for better AMS and emphasizing very low use of this free online course among practicing doctors’ community, needs more awareness. Fair theoretical knowledge is seen among doctors with more than fifty percent near correct responses except for the question asking IV route of AM administration which suggests to initiate urgently low-hanging fruits like IV to oral switch or STOP order in the hospital. The surgeon needs more awareness and training on the right dose and spectrum of AM to consider. A significant knowledge gap is found in comparison among faculty, SRs, and JRs reflecting faculty should be at the front end to decide on AM prescriptions rather than free hands residents. Almost all the participants agree that AMS is a necessity in their hospital and believed that it reduces healthcare costs and adverse effects of inappropriate AM prescription. Hence, education on AMS courses is the key to prevent AMR.

## Data Availability

Data is available with the PI.

## Acknowledgements

Ms Anjali Chauhan helped in drafting the data.

## Transparency declarations

None

## Ethical approval

The study was approved by the Institutional Ethics Committee, All India Institute of Medical Sciences, Rishikesh, India under short term studentship program by Indian Council of Medical Research, India (Reference id: 2019-03933).

